# Impact of Total Hip Replacements on the Incidence of Hip Fractures in Norway During 1999–2019. A NOREPOS Study

**DOI:** 10.1101/2022.03.29.22272896

**Authors:** Helena Kames Kjeldgaard, Haakon E Meyer, Martin O’Flaherty, Ellen M Apalset, Cecilie Dahl, Nina Emaus, Anne Marie Fenstad, Ove Furnes, Jan-Erik Gjertsen, Mari Hoff, Berit Schei, Anne Johanne Søgaard, Grethe S Tell, Kristin Holvik

**Author notes:** Corresponding author: Helena Kames Kjeldgaard, Norwegian Institute of Public Health, Department of Physical Health and Ageing, Marcus Thranes gate 6, 0473 Oslo, Norway.

## Abstract

The knowledge about why hip fracture rates in Norway have declined is sparse. Concurrent with decreasing hip fracture rates, the rates of total hip replacements (THRs) have increased. We wanted to investigate if hip fracture rates continued to decline, and whether the increase in THRs had any influence on this decline, assuming that living with a hip prosthesis precludes fracture of the operated hip. Information on hip fractures in Norway 1999-2019 was available from the Norwegian Epidemiologic Osteoporosis Studies (NOREPOS) hip fracture database and population size were available in official population tables from Statistics Norway. Primary THRs (for any cause except hip fracture) 1989-2019 were obtained from the Norwegian Arthroplasty Register. We calculated the annual age-standardized incidence rates of hip fracture by sex for the period 1999–2019. The hip fracture rates in a scenario with no hip prostheses were calculated by subtracting 0.5 persons from the population at risk for each prevalent hip prosthesis, considering that each person has two hips at risk of fracture. We estimated how much of the decline could be attributed to the increased prevalence of hip prostheses. From 1999 to 2019, age-standardized incidence rates of hip fracture decreased by 27% in women and 20% in men. The rates remained stable in those under 70 years and decreased in those 70 years and above. Excluding replaced hips from the population at risk led to higher incidence rates, and this impact was considerably larger at higher ages. The increased prevalence of hip prostheses over the period accounted for approximately 18% (20% in women and 11% in men) of the observed decline in hip fracture rates. In conclusion, the incidence of hip fractures continued to decline, and the increasing number of people living with hip prostheses contributed significantly to the observed declining time trends.

## INTRODUCTION

Hip fractures among older adults are associated with a high burden of morbidity and mortality.^(1)^ With continuing increasing life expectancy and population growth, the proportion of people aged 60 years and older will increase substantially over the next few decades, ^(2)^ forecasting an increasing societal burden of hip fractures.^(3)^ There is a large variation in hip fracture incidence rates between countries, with the highest rates in Scandinavia. ^(4)^ Decreasing incidence rates have been observed in most Western countries in recent decades,^(5)^ including Norway.^(6)^ The causes for the decrease in hip fracture rates are not clear, although several contributing factors have been suggested.^(6)^ Concurrent with decreasing hip fracture rates, the rates of total hip replacements (THRs) have increased.^(7)^ The dominating indication for a THR is osteoarthritis of the hip, a chronic and progressive condition with loss of cartilage that increases with age.^(8)^ With increasing life expectancy and a growing elderly population, the THR rates are expected to increase further.^(9)^

In Norway, the lifetime risk of primary THR for osteoarthritis was estimated to be 16% for women and 8.3% for men in 2013,^(10)^ and the annual number of primary THRs has constantly increased.^(11)^ Further, the average age when receiving hip replacement has decreased over time.^(11)^

Assuming that a hip replacement practically eliminates the risk of fracturing the operated hip, we hypothesized that the increasing population prevalence of persons living with hip prostheses may have contributed to the observed declining trends in risk of hip fracture in Norway during recent decades.^(6)^

The aims of the present study were to 1) investigate trends in hip fracture incidence rates in Norway from 1999 to 2019 in Norwegians 50 years and older, and 2) explore the degree to which an increase in the prevalence of people living with hip prosthesis has affected the incidence of hip fracture.

## METHODS

### Study Population and Data Sources

The study covered the population of Norway aged 50 years and older in the period 1999 through 2019. Population size as of 1^st^ January for each calendar year 1999 to 2020 according to sex and 1-year age groups was available in official population tables published by Statistics Norway (www.ssb.no/en). Within each sex- and 1-year age group, the midyear population in each calendar year was defined as the arithmetic mean of the number with index age on January 1^st^ of the respective year and the number with index age+1 on January 1^st^ of the subsequent year.

Information on all inpatient contacts with a hip fracture diagnosis in specialist healthcare in Norway 1994-2019 was available in the Norwegian Epidemiologic Osteoporosis Studies (NOREPOS) hip fracture database (NORHip),^(12)^ including cervical, trochanteric or sub-trochanteric fractures (ICD 9: 820 with all subgroups; ICD 10: S72.0– S72.2). Data from 1994 to 2007 were obtained from the patient administrative systems in the treating hospitals, and the corresponding data for 2008 through 2019 was obtained from the Norwegian Patient Registry. Using information about co-occurrence of other diagnosis codes, surgical procedure codes, and whether the hip fracture was registered as a primary or secondary diagnosis, a comprehensive algorithm was applied to identify the hospitalizations that represented a newly occurred (incident) hip fracture. This algorithm has been validated and have shown high agreement with quality-checked data (www.norepos.no/documentation). Up to two hip fractures per person were included, as was done previously.^(6,13)^ Age at discharge was calculated. We included fractures from 1999 onwards in correspondence with previous analyses, allowing a five-year washout period.

Information on primary total hip replacements in Norway during 1989-2019 were obtained from the Norwegian Arthroplasty Register (NAR), established in 1987 and is considered complete from 1989 onwards.^(14)^ NAR is based on individual reporting from each operation conducted by the orthopedic surgeons. Registration in NAR is based on informed consent from the patients. Completeness for primary total hip replacements in NAR has been reported to be 97%.^(11,15)^

### Statistical Analyses

We calculated the incidence rates of hip fracture according to sex and 5-year age groups from 50 to 90+ years in three 7-year periods; 1999-2005, 2006-2012, and 2013-2019. Age-standardization of hip fracture rates was done by the direct method using the population distribution of 1^st^ January 2019, separately for women and men. These rates were used to examine trends in hip fracture incidence from 1999 to 2019.

The number of prevalent hip prostheses in living individuals in 2019 was based on data from NAR in the period 1989-2019. Assuming that a limited number of persons receiving THR before 1989 would be alive by 2019, the 2019 prevalence was estimated by calculating the number of primary THRs for any indication other than hip fracture (acute or sequelae) performed between 1989 and 2019 in individuals who were alive and 50 years or older by 1^st^ January 2019. While the number of prevalent hip prostheses in 2019 was based on 30 years of reporting to NAR (1989-2018), THR data were available for only one decade before 1999. In order to estimate the total number of prevalent hip prostheses per 1^st^ January 1999, we used information from the NAR in the period 1989-1998, a report on THRs in Norway in 1980^(16)^, and discussions with a former Principal Investigator of NAR. The number of THRs performed in 1980 (2,435 primary operations) was approximately half of the mean yearly number of primary operations performed in 1989-1991. We assumed that primary operations had also doubled from 1969 to 1978. Based on this we assumed a linear increase of primary operations (a doubling from 1969 to 1979 and another doubling from 1979 to 1989).

To explore the possible impact of changing population prevalence of hip prostheses on the hip fracture incidence, we estimated the hip fracture incidence in a scenario with no prevalent hip prostheses in the population at risk. In this scenario, 0.5 persons were subtracted from the population at risk for each prevalent hip prosthesis, considering that each person has two hips at risk of fracture and that a person with one hip has a relative risk (RR) of 0.5 for a new hip fracture compared to a person with two hips. We verified this assumption in an additional analysis in individual-level data on all inhabitants aged 50-80 years in 2002 in the Norwegian Population and Housing Census 2001 linked with data from NAR and NORHip to identify primary THR and hip fractures during 2002-2018. We excluded patients with THR before 2002 (15 years’ washout). The association between THR and risk of hip fracture was estimated using Cox proportional hazards regression with attained age as timescale, with THR as time-dependent exposure. All inhabitants aged 50-80 years without previous THR were followed until hip fracture (event) or censoring due to a second hip replacement, emigration, death, or 31^st^ December 2018, whichever occurred first.

In addition, we estimated how much of the decline in hip fracture incidence rate could be attributed to increase in the prevalence of hip prostheses in the population. First, we calculated the expected number of hip fractures in 2019 given unchanged age- and sex specific hip fracture rates since 1999. We then calculated the difference between the observed and expected number of hip fractures in 2019. The difference that could be attributed to increased prevalence of hip prostheses in the population was estimated as the difference between the population attributable fractions (PAF) in 1999 and 2019, multiplied by the expected number of hip fractures in 2019, where PAF = (prevalence of hip prostheses*(RR-1)/ (prevalence of hip prostheses*(RR-1) +1) and the RR of hip fractures was set to 0.5.

Using Monte Carlo simulation, we calculated 95% uncertainty intervals for the difference in expected and observed number of hip fractures that was attributed to hip prostheses.^(17)^ This was done by replacing the input parameters by appropriate probability distributions and repeatedly recalculating the output with values randomly sampled from the defined input distributions. We used the Excel add-in Ersatz software (version 1.35 available at www.epigear.com) to perform 1000 runs to determine the 95% uncertainty intervals of the hip fractures prevented (2.5th and 97.5th percentile values corresponding to the lower and upper limits, see Supplemental Table 1).

### Ethics

The study and the linkages of data were approved by the Regional Committee for Medical and Health Research Ethics (REK South-East A, ref 15538), the Norwegian Directorate of Health (The Norwegian Patient Registry), the Norwegian Institute of Public Health and Statistics Norway. The data were handled in accordance with the General Data Protection Regulation, and a Data Protection Impact Assessment has been conducted.

## RESULTS

From 1999 to 2019 the annual number of hip fractures in women decreased 11% (from 6,749 to 6,037), while it increased 20% in men (from 2,560 to 3,084). However, the age-standardized hip fracture rate decreased in both women and men over the period. Women’s age-standardized hip fracture rate decreased from 88 per 10,000 in 1999 to 64 per 10,000 in 2019. For men, the hip fracture rate per 10,000 decreased from 44 in 1999 to 35 in 2019 (Figure 1, Table 1). These rates correspond to a 27% decline in women and a 20 % decline in men. The female to male hip fracture rate ratio also declined from 2 in 1999 to 1.8 in 2019.

**Table 1.** Total annual number of hip fractures and age-standardized hip fracture incidence rates per 10,000 person-years in Norwegian women and men aged ≥ 50 years from 1999 to 2019.

| Year | Women |  |  |  |  | Men |  |  |  |  |
| --- | --- | --- | --- | --- | --- | --- | --- | --- | --- | --- |
|  | Number of fractures<sup>a</sup> | Population<sup>b</sup> | Crude rate<sup>c</sup> | Standardized rate<sup>d</sup> | 95% CI | Number of fractures<sup>a</sup> | Population<sup>b</sup> | Crude rate<sup>c</sup> | Standardized rate<sup>d</sup> | 95% CI |
| 1999 | 6749 | 739638 | 91 | 88 | 86, 90 | 2560 | 626707 | 41 | 44 | 42, 45 |
| 2000 | 6596 | 747521 | 88 | 85 | 83, 87 | 2465 | 637261 | 39 | 41 | 40, 43 |
| 2001 | 6779 | 755060 | 90 | 86 | 84, 88 | 2595 | 647577 | 40 | 43 | 41, 45 |
| 2002 | 6671 | 761613 | 88 | 84 | 82, 86 | 2490 | 657223 | 38 | 40 | 39, 42 |
| 2003 | 6767 | 769694 | 88 | 84 | 82, 86 | 2594 | 668232 | 39 | 42 | 40, 43 |
| 2004 | 6786 | 779128 | 87 | 83 | 81, 85 | 2599 | 680070 | 38 | 41 | 40, 43 |
| 2005 | 6588 | 789171 | 83 | 80 | 78, 82 | 2584 | 692451 | 37 | 40 | 39, 42 |
| 2006 | 6651 | 799555 | 83 | 80 | 78, 82 | 2770 | 705926 | 39 | 42 | 41, 44 |
| 2007 | 6750 | 810503 | 83 | 80 | 79, 82 | 2748 | 720209 | 38 | 41 | 39, 43 |
| 2008 | 6725 | 821206 | 82 | 80 | 78, 82 | 2865 | 734266 | 39 | 42 | 41, 44 |
| 2009 | 6567 | 832638 | 79 | 77 | 75, 78 | 2746 | 749047 | 37 | 39 | 38, 41 |
| 2010 | 6480 | 844164 | 77 | 75 | 73, 77 | 2835 | 764520 | 37 | 40 | 39, 42 |
| 2011 | 6556 | 855632 | 77 | 75 | 74, 77 | 2932 | 779973 | 38 | 40 | 39, 42 |
| 2012 | 6555 | 867259 | 76 | 75 | 73, 76 | 2832 | 795838 | 36 | 38 | 37, 40 |
| 2013 | 6575 | 879306 | 75 | 75 | 73, 76 | 2910 | 811920 | 36 | 38 | 37, 40 |
| 2014 | 6389 | 893025 | 72 | 71 | 70, 73 | 2773 | 828527 | 33 | 36 | 34, 37 |
| 2015 | 6208 | 907892 | 68 | 69 | 67, 71 | 2946 | 846920 | 35 | 37 | 35, 38 |
| 2016 | 6306 | 923106 | 68 | 69 | 68, 71 | 2921 | 865400 | 34 | 36 | 34, 37 |
| 2017 | 6095 | 938734 | 65 | 66 | 65, 68 | 3056 | 884296 | 35 | 36 | 35, 38 |
| 2018 | 6114 | 954851 | 64 | 66 | 64, 67 | 3038 | 903065 | 34 | 35 | 34, 36 |
| 2019 | 6037 | 972093 | 62 | 64 | 62, 66 | 3084 | 923145 | 33 | 35 | 33, 36 |

CI confidence interval
a. Total number of hip fractures in people $\geq 50$ years in the NORHip database
b. Calculated mid-year population
c. Crude hip fracture rate per 10,000 person-years
d. Age-standardized hip fracture rate per 10,000 person-years. The standard population was the population distribution of 1<sup>st</sup> January 2019, separately for women and men

**Figure 1.**
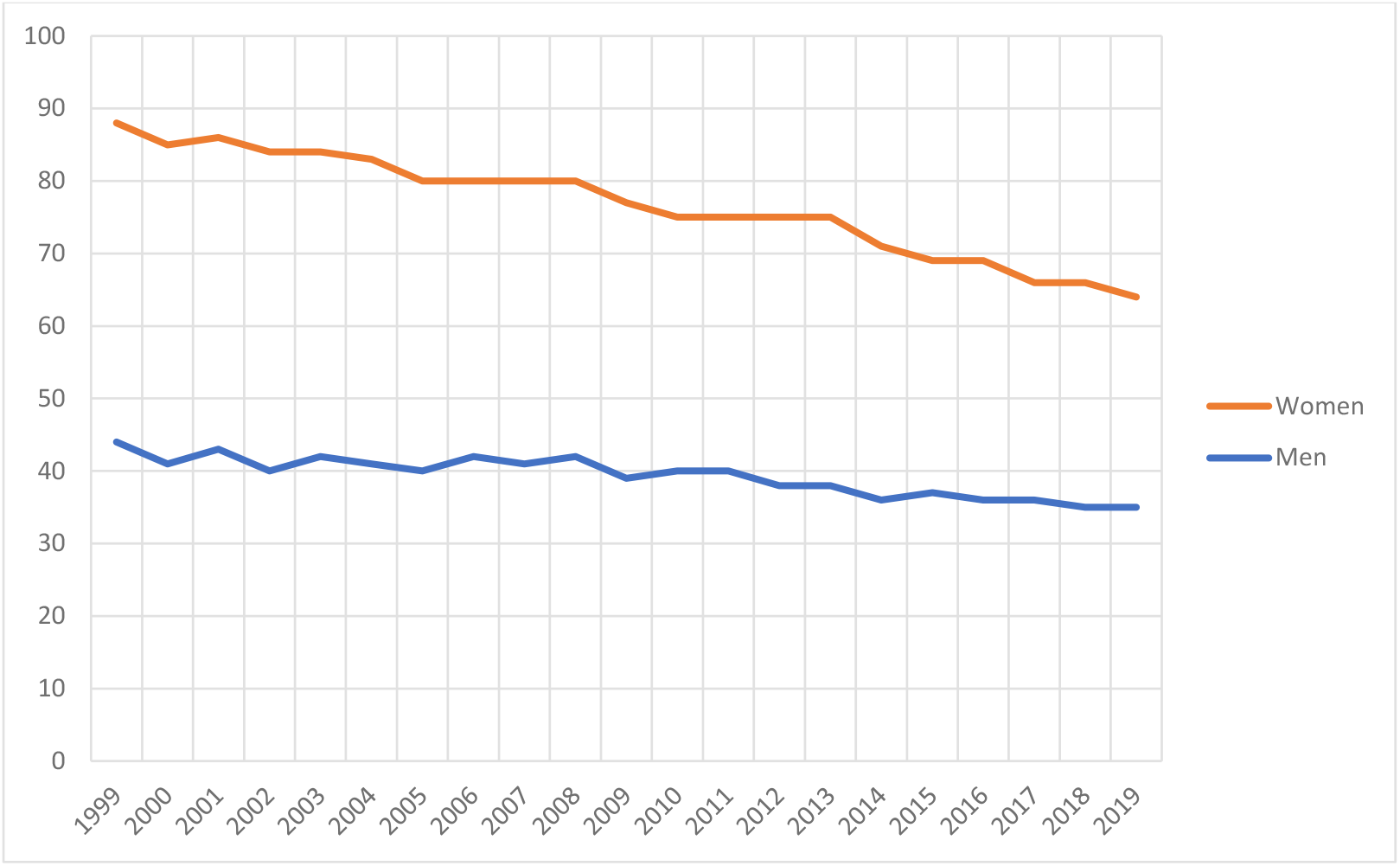
Age-standardized hip fracture incidence rates per 10,000 person-years in women and men from 1999 to 2019.

For both women and men under 70 years, the incidence rate remained stable over the three time periods. For women and men aged 70 and above, the hip fracture rates decreased over the three time periods, with the only exception being men aged 90 and above who had the highest incidence in 2006-2012 (Table 2).

**Table 2.** Number and incidence rates per 10,000 person-years of hip fractures in 5-year age groups (≥ 50 years) in the Norwegian population. Data from women and men in the three 7-year time periods 1999-2005, 2006-2012, and 2013-2019.

| Age | 1999-2005 |  |  | 2006-2012 |  |  | 2013-2019 |  |  |
| --- | --- | --- | --- | --- | --- | --- | --- | --- | --- |
|  | Number of hip fractures<sup>a</sup> | Population<sup>b</sup> | Incidence per 10,000 | Number of hip fractures<sup>a</sup> | Population<sup>b</sup> | Incidence per 10,000 | Number of hip fractures<sup>a</sup> | Population<sup>b</sup> | Incidence per 10,000 |
| Women |  |  |  |  |  |  |  |  |  |
| 50-54 | 505 | 1034860 | 5 | 478 | 1079847 | 4 | 423 | 1173529 | 4 |
| 55-59 | 819 | 914195 | 9 | 962 | 1022575 | 9 | 907 | 1089860 | 8 |
| 60-64 | 1167 | 692201 | 17 | 1500 | 957696 | 16 | 1625 | 1015633 | 16 |
| 65-69 | 1910 | 608017 | 31 | 2162 | 727210 | 30 | 2827 | 954999 | 30 |
| 70-74 | 3756 | 607455 | 62 | 3262 | 575826 | 57 | 4010 | 766869 | 52 |
| 75-79 | 7571 | 599553 | 126 | 5832 | 520751 | 112 | 5737 | 544293 | 105 |
| 80-84 | 11685 | 483931 | 241 | 9884 | 459320 | 215 | 8258 | 420558 | 196 |
| 85-89 | 11431 | 274994 | 416 | 12099 | 323039 | 375 | 10215 | 304242 | 336 |
| 90+ | 8092 | 126541 | 639 | 10105 | 164591 | 614 | 9722 | 198916 | 489 |
| Men |  |  |  |  |  |  |  |  |  |
| 50-54 | 482 | 1078226 | 4 | 510 | 1118437 | 5 | 539 | 1238091 | 4 |
| 55-59 | 653 | 935146 | 7 | 761 | 1053518 | 7 | 781 | 1126384 | 7 |
| 60-64 | 827 | 671731 | 12 | 1206 | 970076 | 12 | 1254 | 1026559 | 12 |
| 65-69 | 1067 | 551512 | 19 | 1381 | 696033 | 20 | 1842 | 947543 | 19 |
| 70-74 | 1850 | 504742 | 37 | 1789 | 501529 | 36 | 2439 | 719372 | 34 |
| 75-79 | 3281 | 428670 | 77 | 2747 | 404548 | 68 | 2995 | 456795 | 66 |
| 80-84 | 4198 | 277454 | 151 | 4129 | 295660 | 140 | 3757 | 301504 | 125 |
| 85-89 | 3492 | 121637 | 287 | 4361 | 156392 | 279 | 3991 | 172396 | 232 |
| 90+ | 2037 | 40310 | 505 | 2844 | 53492 | 532 | 3130 | 74529 | 420 |

a. Total number of hip fractures in people $\geq 50$ years in the NORHip database
b. Calculated mid-year population

Concurrent with the declining hip fracture trend, the number of people living with hip prostheses increased. As shown in Table 3, the number of people living with at total hip prosthesis roughly doubled in all age groups in both women and men from 1999 to 2019. In 2019, 2,756 fewer hip fractures were observed (N = 9,121) than expected (N = 11,877) if the hip fracture rates had remained constant since 1999.

**Table 3.** Hip fracture incidence rates in three age groups for Norwegian women and men ≥ 50 years observed in 1999 and 2019 and calculated in a scenario with no hip prostheses in the population at risk.

| Age | Number of hip fractures<sup>a</sup> | Number of hip prostheses<sup>b</sup> | Population<sup>c</sup> | Observed hip fracture incidence<sup>d</sup> | Fracture incidence in scenario<sup>e</sup> |
| --- | --- | --- | --- | --- | --- |
| Women 1999 |  |  |  |  |  |
| 50-64 | 334 | 4251 | 347463 | 10 | 10 |
| 65-79 | 2121 | 18909 | 273054 | 78 | 80 |
| 80+ | 4294 | 14957 | 119113 | 361 | 385 |
| Women 2019 |  |  |  |  |  |
| 50-64 | 413 | 8984 | 488159 | 8 | 9 |
| 65-79 | 1923 | 32551 | 352006 | 55 | 57 |
| 80+ | 3701 | 31086 | 131919 | 281 | 318 |
| Men 1999 |  |  |  |  |  |
| 50-64 | 254 | 2440 | 349772 | 7 | 7 |
| 65-79 | 943 | 8098 | 218917 | 43 | 44 |
| 80+ | 1363 | 4611 | 58006 | 235 | 245 |
| Men 2019 |  |  |  |  |  |
| 50-64 | 347 | 6700 | 505563 | 7 | 7 |
| 65-79 | 1164 | 17851 | 334738 | 35 | 36 |
| 80+ | 1073 | 10671 | 82833 | 130 | 138 |

a. Number of hip fractures in the NORHip database
b. Number of prevalent hip prostheses in living individuals based on information from the Norwegian Arthroplasty Register
c. Calculated midyear population
d. Observed hip fracture incidence per 10,000
e. The hip fracture incidence rates per 10,000 in a scenario with no hip prostheses were calculated by subtracting 0.5 persons from the population at risk for each prevalent hip prosthesis

We estimated that 18% (20% in women and 11% in men) of the difference between the observed and expected number of hip fractures in 2019 could be attributed to the increased prevalence of persons living with a total hip prosthesis (Figure 2). As shown in the Supplemental Table 2, the 95% uncertainty intervals were narrow. In a sensitivity analysis allowing the prevalence estimate in 1999 to vary by 20% in each direction only increased the uncertainty interval marginally (from 17-18% to 16-19%).

**Figure 2.**
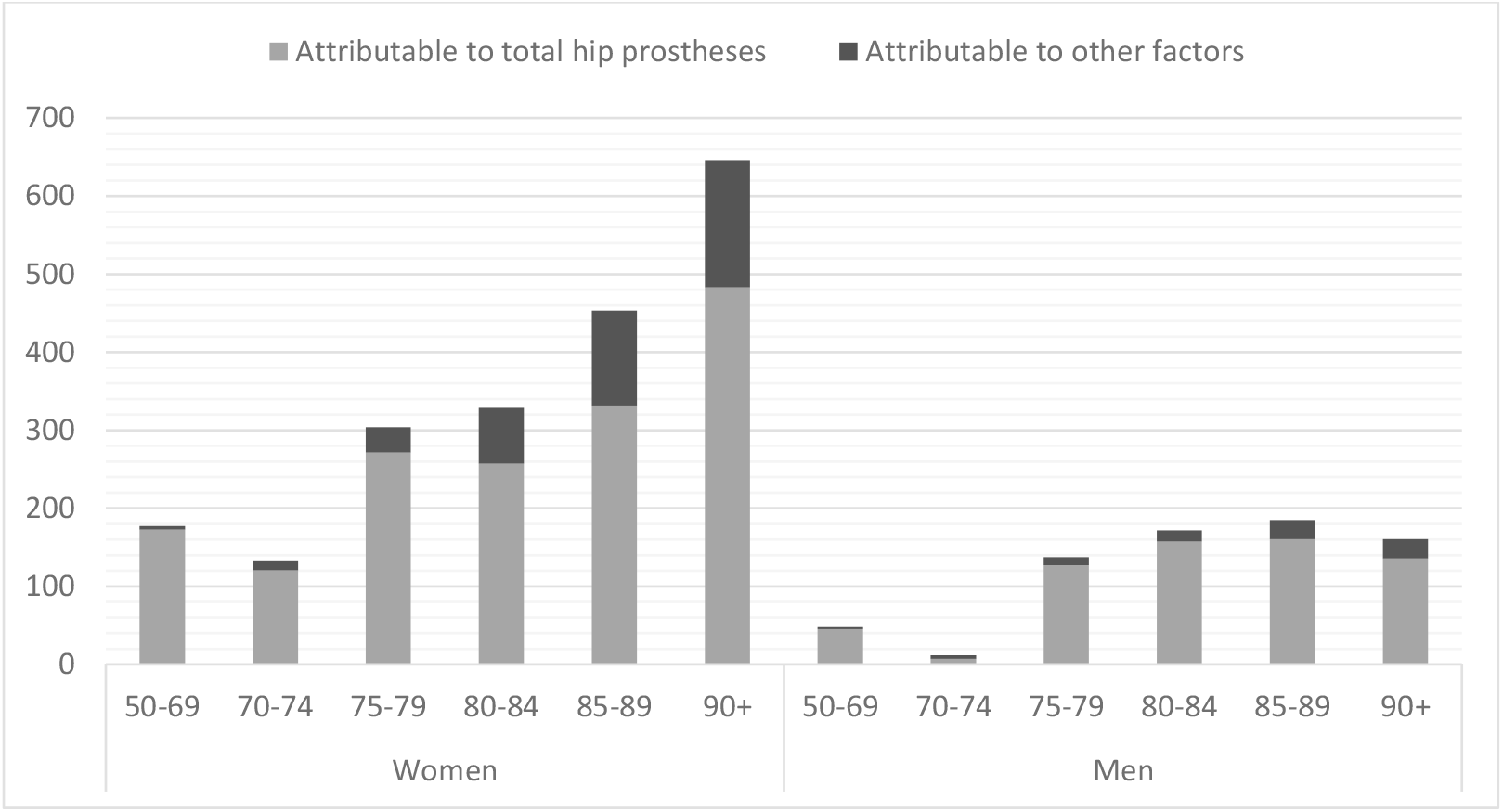
The total column heights represent the difference between the observed and expected number of hip fractures in six age groups in women and men in 2019 given unchanged hip fracture rates since 1999. The dark parts of the columns represent the differences attributable to the increased prevalence of hip prostheses from 1999-2019.

In these calculations we assumed that a total hip replacement is associated with a RR of 0.5 for a subsequent fracture in the un-operated hip. This was confirmed in our additional analysis on 1,268,110 individuals aged 50-80 years with 65,772 primary total hip replacements and 93,882 hip fractures during up to 17 years follow-up. The relative risk of hip fracture after having a THR was 0.50 (95% CI 0.48, 0.52) compared to not having a THR.

## DISCUSSION

It has previously been reported that the incidence of hip fracture in Norway declined from 1999 to 2013,^(6)^ and the current report shows that this decline in age-adjusted rates continued in the period 2014-2019. Hip fracture rates declined significantly in almost all 5-year age groups among those 70 years and older. The decrease in incidence rates were larger in women compared to men and the female/male hip fracture rate ratio was slightly reduced. However, due to demographic changes, the number of hip fractures in the population has been fairly stable with 9,309 fractures in 1999 and 9,121 fractures in 2019.

In recent decades, decreasing age-adjusted hip fracture incidence rates have been reported in most Western countries.^(5)^ As discussed in an earlier NOREPOS study,^(6)^ several contributing factors for this decline have been suggested such as osteoporosis medication, reduced smoking, increased BMI, and increased physical activity; however overall, the causes for this decline are not clear. Using a novel approach, we explored whether the increased use of total hip replacements could explain part of the declining hip fracture rates. Creating a scenario with no hip prostheses in the population at risk, we found that hip fracture incidence rates were overall higher compared to the observed incidence rates, notably in those over 80 years. The results indicated that about 18% of the decline in hip fractures rates from 1999 to 2019 could be attributed to the increased prevalence of people living with hip prostheses.

In Norway, the primary indication of total hip replacements is osteoarthritis. It is unclear how osteoarthritis influences the hip fracture risk.^(18)^ However, a high body mass index is a strong risk factor for osteoarthritis of the hip^(19)^ while reducing hip fracture risk.^(20)^ In the present study we found that a total hip replacement was associated with a RR of 0.5 for a subsequent fracture in the un-operated hip, confirming our assumption that a person with one un-operated hip has a RR of 0.5 for a new hip fracture compared to a person with two intact hips.

Despite that hip fracture rates continued to decline through 2019, further actions are most probably needed to counteract a rise in the annual number of hip fractures as the number of older adults is increasing. According to projections by Statistics Norway, the number of people aged 70 and over will nearly double from 2020 to 2050 (www.ssb.no). The average annual decline of hip fractures in women between 1999-2019 was 1.3%, however, earlier projections of the burden of hip fractures in Norwegian postmenopausal women suggest that an annual decrease of this magnitude until 2040 would not be sufficient to counteract the effects of the ageing population.^(3)^ An increasing number of immigrants from populations with lower hip fracture rates may affect future hip fracture trends in Norway. However, from 1999-2019 this immigrant group was relatively young and is unlikely to explain the reduction in hip fracture rates.^(21)^

A strength of this study was the inclusion of national data on hip fractures by retrieving information on all cervical, trochanteric, or subtrochanteric hip fractures treated in Norwegian hospitals in the period 1999-2019 available in the NORHip database. Norway has a universal health coverage, and the completeness and validity of this database has been documented (www.norepos.no/documentation).

NAR has complete national coverage and the completeness of primary total hip replacements has been found to be 97.5%. This completeness has been stable over the years.^(11,15)^ A limitation was lacking individual information on THR before NAR gained complete national coverage in 1989, which could impact on our calculation of how much of the decline could be attributed to THRs. However, sensitivity analyses suggest that these calculations are robust.

In conclusion, the age-standardized incidence of hip fractures declined by 27% in women and 20% in men from 1999 to 2019. The increasing number of people living with hip prostheses explained nearly one-fifth of this decline.

## Data Availability

The current study is part of a research project at the Norwegian Institute of Public Health. The datasets generated and analyzed during this study are not publicly available due to data protection regulations but are made available to project members on a remote access platform. Alternatively, these data can be reconstructed anew by applying to each registry.

## ACKNOWLEDGEMENTS

All authors state no conflict of interest.

We thank the Norwegian orthopedic surgeons for conscientiously reporting all their THR operations and the staff at the Norwegian Arthroplasty Register for entering the data and statistical advice.

## Disclaimer

Data from the Norwegian Patient Registry have been used in this publication. The interpretation and reporting of these data are the sole responsibility of the authors, and no endorsement by the Norwegian Directorate of Health is intended nor should be inferred.

## Notes

### Competing Interest Statement

The authors have declared no competing interest.

### Funding Statement

This study received funding from the Research Council of Norway (grant number 275270)

