## Supplemental tables for "Impact of Total Hip Replacements on the Incidence of Hip Fractures in Norway During 1999–2019. A NOREPOS Study"

**Supplemental Table 1.** Uncertainty analysis using Monte Carlo simulation Excel add-in Ersatz software.

| Parameter | Distribution | Ersatz function | Source |
|---|---|---|---|
| Population | Poisson | ErPoisson | Statistics Norway |
| Hip fractures | Poisson | ErPoisson | NOREPOS hip fracture database |
| Total hip replacement proportion | Beta | ErBeta | The Norwegian Arthroplasty Register |
| Relative risk | Lognormal | ErRR | Results presented in this paper |

**Supplemental Table 2.** The difference between expected and observed number of hip fractures in Norway in 2019 and the difference attributable to the increased prevalence of hip prostheses from 1999-2019.

| Age | Difference between expected and observed number of hip fractures in 2019[a] | Difference attributed to hip prostheses, N | Difference attributed to hip prostheses, % |
|---|---|---|---|
| Women | | | |
| 50-54 | 21 | 0 | - |
| 55-59 | 9 | 0 | - |
| 60-64 | 63 | 1 | 2 % |
| 65-69 | 84 | 3 | 4 % |
| 70-74 | 133 | 13 | 9 % |
| 75-79 | 304 | 33 | 11 % |
| 80-84 | 329 | 72 | 22 % |
| 85-89 | 453 | 122 | 27 % |
| 90+ | 646 | 163 | 25 % |
| Total | 2042 | 406 (391, 422)[b] | 20 % (19, 21)[b] |
| Men | | | |
| 50-54 | 14 | 0 | - |
| 55-59 | 14 | 0 | - |
| 60-64 | 16 | 1 | 5 % |
| 65-69 | 4 | 1 | 34 % |
| 70-74 | 12 | 5 | 39 % |
| 75-79 | 137 | 10 | 7 % |
| 80-84 | 171 | 14 | 8 % |
| 85-89 | 185 | 24 | 13 % |
| 90+ | 160 | 25 | 15 % |
| Total | 713 | 80 (75, 85)[b] | 11 % (11, 12)[b] |
| Women and men total | 2756 | 486 (469, 502)[b] | 18 % (17, 18)[b] |

a. The difference between the observed and expected number of hip fractures in 2019 given unchanged hip fracture rates since 1999

b. Uncertainty interval calculated using Monte Carlo simulation
